# Challenges faced by Human Resources for Health in Morocco: a scoping review

**DOI:** 10.1101/2023.12.21.23300411

**Authors:** Wafaa Al Hassani, El Youness Achhab, Chakib Nejjari

## Abstract

**Background:** Human resources for health (HRH) play a pivotal role in effective health system operation, yet various impediments challenge sustainable development. This scoping review aimed to explore these challenges and potential solutions in aligning the health workforce to meet the evolving healthcare needs of the Moroccan population.

**Methods:** We conducted a scoping review searching PubMed, Science Direct, Cairn and Google Scholar for relevant articles published between 2014 and 2023. Additionally, non-peer-reviewed literature sourced from Ministry of Health consultations and allied websites was included.

**Results:** Among the nineteen studies meeting our inclusion criteria, the majority were cross-sectional and predominantly focused on challenges faced by nurses. While some papers delineated multiple HRH challenges (5/19), the rest addressed specific challenges. The identified challenges span organizational and personal levels. Organizationally, the focus was on training, lifelong learning, continuing education, health coverage and shortages, and job satisfaction. At a personal level, HRH in the public health sector encountered challenges such as burnout, stress, and broader occupational health concerns.

**Conclusions:** The reviewed publications underscored a spectrum of challenges necessitating robust policy interventions. Despite promising developments in the Moroccan healthcare system, addressing the unequal urban-rural HRH distribution, augmenting funding, and enhancing HRH quality of life stand as pivotal imperatives.

## Introduction

Human resources for health (HRH) serve as a linchpin for robust health systems worldwide, yet multiple challenges impede their sustainable development [1–5]. In 2016, the World Health Assembly introduced the Global Strategy on Human Resources for Health: Workforce 2030, aimed at expanding the availability, accessibility, acceptability, coverage, and quality of HRH. This strategy stands as a critical step toward achieving universal health coverage and the Sustainable Development Goals [5].

Recent progress in research has delved into various facets of HRH, examining governance, training, performance, recruitment strategies, motivation, and distribution [6–11]. Systematic reviews play a pivotal role in visualizing this evidence for policymakers. They consolidate research, shedding light on key issues affecting the health workforce within a robust health system [12–13]. Such reviews not only highlight areas with existing evidence but also pinpoint those necessitating more attention. Ultimately, fostering a synergistic interaction between researchers and policymakers through these systematic reviews facilitates the translation process within HRH policymaking. This collaborative approach aims to bridge the gap between research insights and policy implementation.

Morocco’s healthcare system is integral to the well-being of its populace, yet it grapples with a myriad of challenges, as do many nations [14]. Despite notable strides in recent years, this system confronts ongoing hurdles that demand readiness from healthcare professionals. The Human Resources for Health (HRH) sector stands as a critical pillar within this system, encompassing healthcare professionals, support staff, and administrators. The Joint Learning Initiative on HRH emphasizes the significance of investing in HRH as a means to explicitly elevate national income per capita and alleviate absolute poverty [15]. Moreover, participants from diverse backgrounds at the fifth Global Forum on HRH in 2023 underscored the imperative of safeguarding, protecting, and investing in the health and care workforce [16].

This paper delineates the outcomes of an initiative aimed at investigating challenges and potential solutions to optimize the health workforce. Through an extensive literature review, the objective was to align the health workforce with the evolving healthcare requirements of the Moroccan population.

## Methods

### Data sources and search strategy

Adhering to the framework outlined by Arksey and O’Malley [17], as updated by Levac et al. [18], we undertook a scoping review of the literature. Our aim was to delve into the challenges and potential solutions concerning the optimization of the health workforce in Morocco. Initially, the research question was formulated through consultations with an expert panel comprising members from the health ministry, an EMRO representative, and a researcher equipped with formal information management training. To establish relevant search criteria, keywords were identified based on these research inquiries and insights provided by the expert panel. Furthermore, the summary and reporting of this scoping review were done using the checklist for the Preferred Reporting Items for Systematic Reviews and Meta-Analyses extension for Scoping Reviews (PRISMA-ScR) [19]. Ethical approval was not deemed necessary for this review since the findings were derived from existing and publicly available literature.

We conducted searches on electronic databases including PubMed, ScienceDirect, and Google Scholar up to November 2, 2023, to identify pertinent articles published between 2014 and 2023. Our search encompassed English and French articles, reviews, abstracts, case reports, letters to the editor, and other academic reports. Additionally, to augment our search, we performed a backward citation analysis of relevant papers and incorporated recommendations provided by our expert panel.

The following key words were identified and used in various combinations with Boolean operators (and, or): health human resources, health care delivery, health workforce, health professionals, healthcare providers, personnel, nurses, doctors, shortage, challenges, barriers, opportunities, satisfaction, stress, burnout, occupational health, Morocco.

A three-stage method was adopted to select publications for review. In the first, the title alone was examined, followed by looking at the abstract, and then examining the whole publication. The search and review results are shown in Fig 1. Articles were excluded if they did not refer to a HRH, or if did not address a challenge context. All documents relevant to the COVID-19 pandemic were excluded from this scoping review. If articles were representative of the inclusion criteria, the documents went through two full-text independent reviews by the two authors.

**Fig 1.**
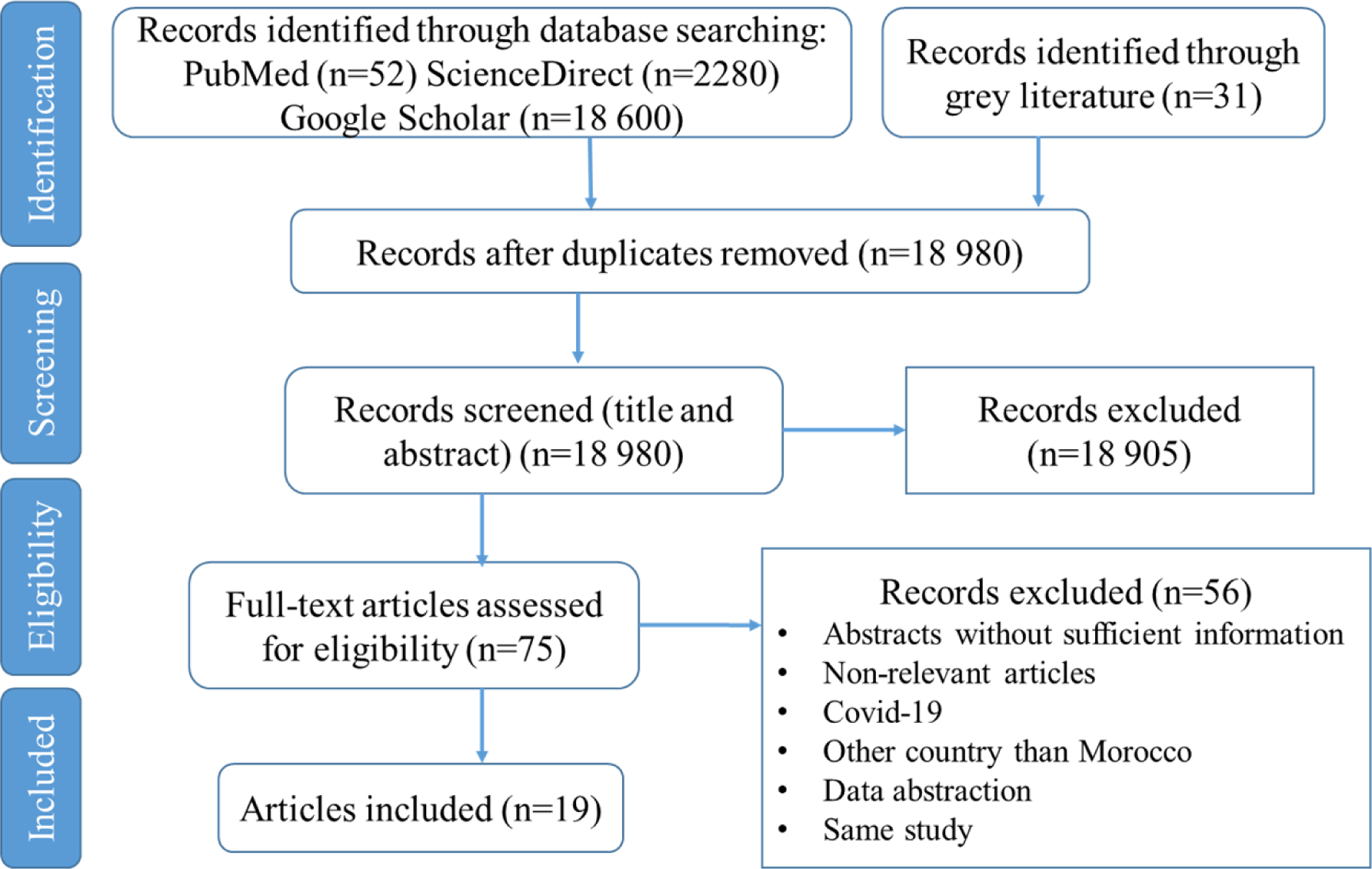
PRISMA flowchart for the study selection process.

### Quality assessment

To assess the literature quality, parts of the method presented by Hölbl *et al*. [20] were used and modified as appropriate. The reviewers checked three considerations about relevance to HRH and challenges and appropriateness of research objectives.

### Data extraction

The research team extracted data for each selected article according to the following domains: author(s) name, year of publication, study design and sample size, type of HRH, type of document (peer-reviewed or gray), setting, HRH challenge, main findings, and proposed solutions or recommendations. Data analysis was completed in Microsoft Excel.

## Results

The selection process is shown in Fig 1. Initially, 18,980 titles were identified through database searches and other sources. Following the screening of titles and abstracts, 75 studies underwent full-text review. Ultimately, 19 studies met the criteria for inclusion in this scoping review. These studies spanned a 10-year range, with the earliest publication dating back to 2014. Interest in challenges encountered by healthcare providers notably surged between 2017 and 2023.

The data extracted from the included studies are presented in Table 1. The majority of publications (15 out of 19) underwent peer review. Among the included studies, half were designed as quantitative cross-sectional (9 out of 19), while four studies adopted a mixed-method approach. Primarily, the focus of most studies was on regional challenges (13 out of 19), with four reports and two articles examining challenges at a national level. However, four publications did not specify the region of their studies. While the majority of studies reported challenges faced by medical professionals, paramedics, and assistants, four studies specifically addressed challenges within one target population, namely nurses.

**Table 1.**
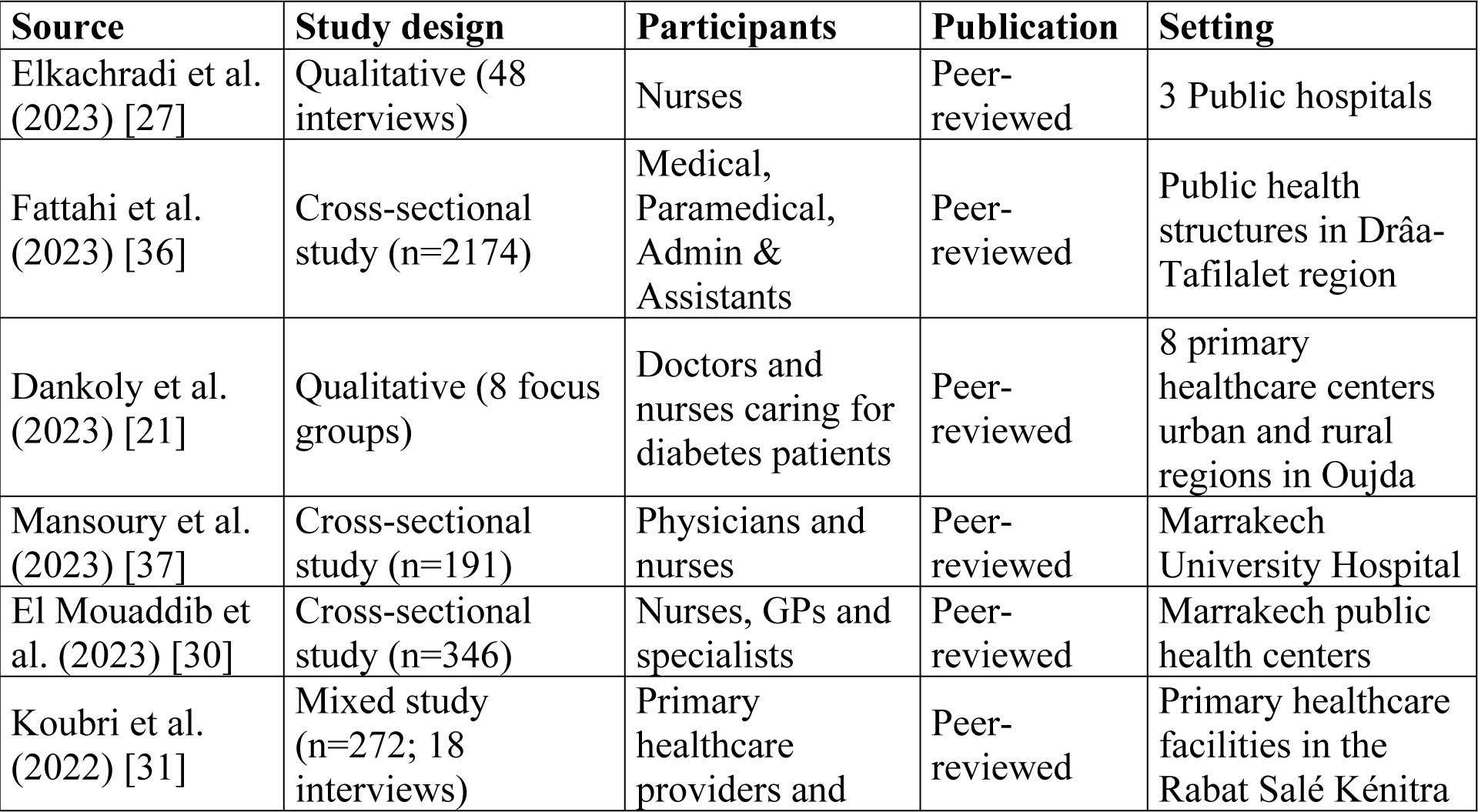
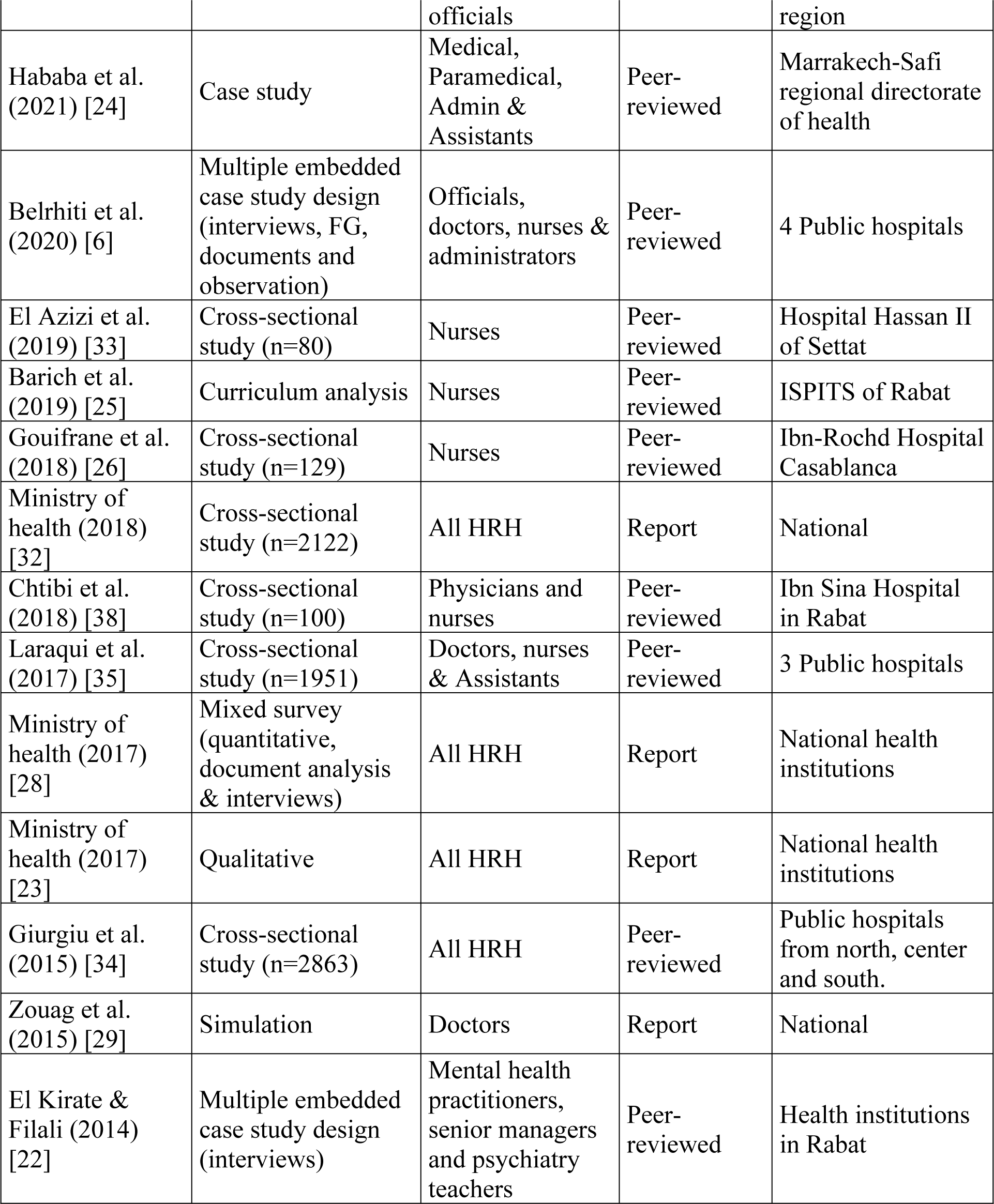
Characteristics of studies selected for inclusion.

| Source | Study design | Participants | Publication | Setting |
| --- | --- | --- | --- | --- |
| Elkachradi et al. (2023) [27] | Qualitative (48 interviews) | Nurses | Peer-reviewed | 3 Public hospitals |
| Fattahi et al. (2023) [36] | Cross-sectional study (n=2174) | Medical, Paramedical, Admin & Assistants | Peer-reviewed | Public health structures in Drâa-Tafilalet region |
| Dankoly et al. (2023) [21] | Qualitative (8 focus groups) | Doctors and nurses caring for diabetes patients | Peer-reviewed | 8 primary healthcare centers urban and rural regions in Oujda |
| Mansoury et al. (2023) [37] | Cross-sectional study (n=191) | Physicians and nurses | Peer-reviewed | Marrakech University Hospital |
| El Mouaddib et al. (2023) [30] | Cross-sectional study (n=346) | Nurses, GPs and specialists | Peer-reviewed | Marrakech public health centers |
| Koubri et al. (2022) [31] | Mixed study (n=272; 18 interviews) | Primary healthcare providers and | Peer-reviewed | Primary healthcare facilities in the Rabat Salé Kénitra |

|  |  | officials |  | region |
| --- | --- | --- | --- | --- |
| Hababa et al. (2021) [24] | Case study | Medical, Paramedical, Admin & Assistants | Peer-reviewed | Marrakech-Safi regional directorate of health |
| Belrhiti et al. (2020) [6] | Multiple embedded case study design (interviews, FG, documents and observation) | Officials, doctors, nurses & administrators | Peer-reviewed | 4 Public hospitals |
| El Azizi et al. (2019) [33] | Cross-sectional study (n=80) | Nurses | Peer-reviewed | Hospital Hassan II of Settat |
| Barich et al. (2019) [25] | Curriculum analysis | Nurses | Peer-reviewed | ISPITS of Rabat |
| Gouifrane et al. (2018) [26] | Cross-sectional study (n=129) | Nurses | Peer-reviewed | Ibn-Rochd Hospital Casablanca |
| Ministry of health (2018) [32] | Cross-sectional study (n=2122) | All HRH | Report | National |
| Chtibi et al. (2018) [38] | Cross-sectional study (n=100) | Physicians and nurses | Peer-reviewed | Ibn Sina Hospital in Rabat |
| Laraqui et al. (2017) [35] | Cross-sectional study (n=1951) | Doctors, nurses & Assistants | Peer-reviewed | 3 Public hospitals |
| Ministry of health (2017) [28] | Mixed survey (quantitative, document analysis & interviews) | All HRH | Report | National health institutions |
| Ministry of health (2017) [23] | Qualitative | All HRH | Report | National health institutions |
| Giurgiu et al. (2015) [34] | Cross-sectional study (n=2863) | All HRH | Peer-reviewed | Public hospitals from north, center and south. |
| Zouag et al. (2015) [29] | Simulation | Doctors | Report | National |
| El Kirate & Filali (2014) [22] | Multiple embedded case study design (interviews) | Mental health practitioners, senior managers and psychiatry teachers | Peer-reviewed | Health institutions in Rabat |

Table 2 outlines the challenges encountered by HRH along with proposed solutions derived from the included studies. While five papers highlighted multiple challenges faced by HRH, the remaining addressed specific challenges. Notably, two papers delineated barriers within distinct departments: the first focused on managing type 2 diabetes [21], while the second discussed challenges within the mental health system [22].

**Table 2.** Distribution of the challenges reported in the includes studies.

| Source | Participants | Challenges | Main results | Proposed solutions |
| --- | --- | --- | --- | --- |
| Elkachradi et al. (2023) [27] | Nurses | Lifelong learning | Only a quarter of the interviewed caregivers comprehend “lifelong learning”. However, all of them acknowledge “continuous learning” particularly amidst prolonged uncertainty, within Moroccan public hospitals. | raise awareness about structuring lifelong professional training while integrating a knowledge management approach |
| Fattahi et al. (2023) [36] | Medical, Paramedical, Admin & Assistants | Burnout syndrome | prevalence rate of burnout (57.7%). Highly stressful working conditions. | Improve working conditions and financial motivation of public health professionals |
| Dankoly et al. (2023) [21] | Doctors and nurses caring for diabetes patients | T2D management barriers | Excessive workload, poor reimbursement policy, shortage, poor working environment, limited referral, gap in the knowledge of general practitioner. | Staff recruitment; continuous professional development; internships. |
| Mansoury et al. (2023) [37] | Physicians & nurses | Burnout syndrome | High degree of emotional exhaustion (60%). High degree of depersonalization. Low degree of personal accomplishment (73%). | Improve working conditions by reduction workload. Increase motivation and lifelong professional training. |
| El Mouaddib et al. (2023) [30] | Nurses, GPs and specialists | Job satisfaction | Presence of dissatisfaction among HCPs regarding various aspects of their jobs. | Clarify the tasks of all professional categories, grant them more autonomy, establish career paths for the promotion and professional development, improve working conditions, and promote incentives. |
| Koubri et al. (2022) [31] | Primary healthcare providers and officials | Satisfaction with organization of health services | Presence of dissatisfaction among HCPs regarding organization of health services. 75% of the providers expressed their full agreement in reorienting PHC | Provision of sufficient and qualified human resources, to be able to develop person-centered health services. Implementing family health as a new paradigm for the |
|  |  |  | facilities towards the person-centered approach. | organization of health services. |
| Hababa et al. (2021) [24] | Medical, Paramedical, Admin & Assistants | Continuing education | Success of the regional continuing education plan for professionals in the Marrakech-Safi region 2019-2021. The participants acquired skills and succeeded in transferring what they acquired in the field. | Developing HRH capacities and improving the performance of the healthcare system. |
| Belrhiti et al. (2020) [6] | Officials, doctors, nurses & administrators | Public service motivation | Complex leadership styles can increase public service motivation, organizational commitment and extra role behaviors by increasing perceived supervisor and organizational support and satisfying staff basic psychological needs. | In hospitals, the archetype of complex professional bureaucracies, leaders need to be able to balance between different leadership styles according to the staff's profile, the nature of tasks and the organizational culture. |
| El Azizi et al. (2019) [33] | Nurses | Evidence-based nursing practice | Organizational barriers, barriers related to nurses' personal characteristics, research barriers, and institutional barriers. | Increase motivation and lifelong professional training. Ensuring access to information sources (scientific databases). |
| Barich et al. (2019) [25] | Nurses | Nursing training | Theoretical courses predominate for all sectors. Objective-based approach instead of skills-based approach. Insufficient time for training. | Need of a second reform referring to the required standards and the specificities of training in nursing and health techniques. |
| Gouifrane et al. (2018) [26] | Nurses | Motivation for continuous training | Level of motivation for continuous training (64%). Extrinsic motivation exceeds intrinsic motivation. | None |
| Ministry of health (2018) [32] | All HRH | Satisfaction, engagement & motivation | Lack of satisfaction (64%). Excess of workload (75%). Stress (75%). 68% unsatisfied | Optimal and appropriate sizing of resources reduces overload and improves |
|  |  |  | from remuneration. 78% satisfied from motivation of their leaderships. Need of continuous training (64%). | the working climate. Ad hoc support structures and innovative solutions contribute to coping with stress. |
| Chtibi et al. (2018) [38] | Physicians & nurses | Stress and Burnout | Emotional exhaustion (42%), depersonalization (49%), low professional achievement (67%), and psychological distress (54%). The stress test reveals that 88% of subjects have a low or moderate level of stress resistance. | None |
| Laraqui et al. (2017) [35] | Doctors, nurses & Assistants | Well-being and occupational perception | Medication use consisted of analgesics (28.1%) and psychotropic (11.6%). Bad general health perception (38.6%). Pain and headache were the most common symptoms. 53.9% experienced stress at work. Difficult work requirements (78.3%). | Assessing the well-being, working conditions and perceived occupational hazards of healthcare staff should be a priority for occupational health departments in Moroccan hospitals. |
| Ministry of health (2017) [28] | All HRH | Continuing training | Diagnosis: Continuing training is very limited, based on an unstructured system and hampered by the lack of funding. | Deploy an efficient and coherent organization for continuing education. strengthen skills to support the Ministry of Health's 2017-2021 sector strategy |
| Ministry of health (2017) [23] | All HRH | Health coverage | Insufficient stock of healthcare workers. Scarce information on dynamics of the healthcare labor market. Maldistribution of healthcare personnel. Insufficient training to guarantee the quality of services. | Increase the availability of HRH according to the service needs. Achieve universal coverage especially in regions with deficits. Design and implement strategies to improve the technical quality of HRH. |
| Giurgiu et al. (2015) [34] | All HRH | Occupational risk perception | Shortage in doctors and nurse staff. Aggression, workload, stress, and musculoskeletal disorders. HCWs took more hypnotics, sedatives and analgesics (40 %). | Urgent need of implementing a risk prevention plan and even a hospital reform. |
| Zouag et al. (2015) [29] | Doctors | Shortage | The baseline scenario shows already important deficits in medical doctors in Morocco for the coming years. This can only increase under the current emigration flows and the retirement rates. | The cooperative frameworks with other countries and mainly EU could be an important source for satisfying both the needs of the EU and those of Morocco. |
| El Kirate & Filali (2014) [22] | Mental health practitioners, senior managers and psychiatry teachers | Challenges faced by the mental health system | Lack of communication, collaboration and updated knowledge especially concerning diagnosis, psychotropic drug prescriptions and addiction medicine. Need of specific training in mental health specialists, such as geriatric psychiatry and pediatric psychiatry. | Better training in these areas would contribute to the success of managed care strategies in primary healthcare facilities. |

Across the included studies, a spectrum of challenges confronted by HRH emerged within both regional and national healthcare sectors. These challenges were categorized into two levels:

### Organizational level

The most frequently reported HRH challenge in the Moroccan context was training, lifelong learning, and continuing education, highlighted in five papers. The Ministry of Health study [23] revealed that continuous training remains significantly limited due to an unstructured system and constrained by insufficient funding. The recommendation from this study emphasizes the necessity of implementing an efficient and coherent organization for continuing education while enhancing the pursuit of quality and attractiveness to strengthen its effectiveness.

Responding to the Ministry of Health’s call for continuous training (outlined in the national strategy of continuous training 2019-2021), the regional directorate of health in Marrakech-Safi devised its regional plan in 2019 [24]. The study demonstrated that participants acquired skills and effectively applied them in practical settings. Among the remaining three papers, all centered on nurses’ training. One study highlighted the need for a secondary reform in the training curriculum [25], while another focused on the inadequacy of motivation for continuous training [26]. Regarding lifelong learning, Elkachradi et al. reported that all healthcare professionals interviewed recognized the importance of training in delivering quality and safe care [27].

Two studies identified health coverage and shortages as key challenges. The first study, conducted by the Ministry of Health [28], highlighted an inadequate stock of healthcare workers that does not align with health service needs. Additionally, it noted an imbalanced distribution of healthcare personnel concerning the geographical spread of the population and its epidemiological profile. The second study, by Zouag et al. [29], focused on forecasting shortages through simulation for the period 2010-2030. It projected a significant deficit in medical doctors in Morocco in the upcoming years. This shortage could further escalate due to current emigration trends and retirement rates.

Three studies shed light on job satisfaction as a noteworthy challenge, characterized by a complex interplay of factors that might vary based on the specific group under study. Overall, dissatisfaction among healthcare providers is prevalent, encompassing various facets of their roles [30–32].

Belrhiti et al.’s study [6] highlights that enhancing public service motivation, organizational commitment, and extra-role behaviors could be achieved by amplifying perceived supervisor and organizational support, along with fulfilling staff’s basic psychological needs. El Azizi and El Goundali’s research [33] focuses on delineating barriers to the integration of evidence-based practices in nursing. It identifies four categories of barriers that could potentially underlie reluctance towards evidence-based practice.

### Personal level

Two studies focused on occupational health highlighted that healthcare workers in Moroccan hospitals tend to use more hypnotics, sedatives, and analgesics [34,35]. Urgent recommendations emerge from these findings, emphasizing the crucial need to assess the well-being, working conditions, and perceived occupational hazards among healthcare staff.

Additionally, two studies underscored the heavy workload experienced within Morocco’s public hospitals, contributing to stress and burnout among healthcare professionals [36,37]. Another study aimed to scrutinize the relationship between resistance status, burnout, and levels of psychological distress [38], revealing that 88% of subjects demonstrated a low or moderate level of stress resistance. Most studies included in this analysis corroborate this stressful scenario and advocate for improving working conditions, as well as promoting incentives and compensation.

## Discussion

This scoping review delved into the challenges confronting HRH within the workforce, as identified in both scientific and grey literature. The study’s robustness lies in its rigorous inclusion criteria and the guidance provided by a panel of experts in shaping the approach to the subject. Notably, nurses emerged as more prominently featured among all professions within the reviewed literature. However, several selected publications encompassed all health professions, primarily due to the comprehensive nature of challenges examined, especially those concerning national health policies.

While a majority of the included studies relied on quantitative methods, the utilization of mixed methods presents an opportunity for triangulating findings. This approach can enrich the contextual understanding of challenges and aid in crafting specific recommendations. Within the body of knowledge on HRH challenges in this scoping review, empirical research takes precedence. Our findings underscore the tendency of empirical studies to concentrate on specific challenges rather than providing a holistic overview. Indeed, the results gleaned from these publications emphasize that challenges are interconnected and often overlap.

Our findings underscore that the most extensively studied challenges since 2014 predominantly align with organizational and personal spheres. Organizational challenges encompass four primary areas: training, lifelong learning, and continuing education; health coverage and shortages; and job satisfaction. Meanwhile, at the personal level, HRH in the public health sector face challenges including burnout, stress, and broader issues related to occupational health. It’s noteworthy that this scenario is not unique to Moroccan HRH but is mirrored in numerous countries worldwide [39–42].

In our digital society, health systems encounter significant challenges, prompting the need to reform models rooted in the industrial age [43]. Responding to a dynamic and complex global environment, the World Health Organization has introduced the ‘Human Resources for Health 2030 strategy’ as a new global paradigm [44]. Central to the realization of this vision is the development of a novel regulatory model. To modernize its regulatory framework, the Moroccan Ministry of Health has devised new strategies in collaboration with the WHO [6,14,45].

The pressing demand to train more healthcare providers to address an estimated shortfall of 18 million, primarily in low- and middle-income countries, presents a critical challenge for health policymakers [5]. One promising strategy to augment the number of medical professionals involves reforming admission and training practices in medical education [46–48]. The focus on training and the future prospects of the health workforce for the 2010-2030 period, as studied by Zouag et al. [29], emphasizes the necessity for enhanced cooperation in healthcare, particularly emphasizing medical education and research. Notably, a study on development assistance for HRH [49] highlighted a significant portion of support directed toward HRH-related training activities between 2016 and 2019.

Following the structuring of continuing training by the Moroccan Ministry of Health [23], local health directorates have devised their strategies. The study conducted by Hababa et al. [24] showcased the effectiveness of a training program executed in 2019 in bolstering HRH skills. Moreover, healthcare professionals view lifelong learning as an ongoing and indispensable process for their work [27,50]. Furthermore, a well-trained HRH is an indispensable element for the success of any healthcare system [51].

This new era is marked by significant transitions, including epidemiologic shifts and a redistribution of the disability burden [4]. These changes significantly impact healthcare systems, the roles of HRH, and educational policies [52,53]. Such transitions further exacerbate the prevalent global shortage and uneven distribution of HRH [1,4].

In Morocco, the Ministry of Health conducted a national study on health coverage [28], revealing an inadequate supply of healthcare workers that doesn’t align with health service needs. Additionally, there’s an imbalanced distribution of healthcare personnel concerning the geographical spread of the population and its health profile. The shortage is compounded by HRH emigration. A recent WHO report indicated that the escalating demand for healthcare providers in developed countries might heighten vulnerabilities in nations already grappling with low health workforce densities [54]. In response, a study in Morocco recommends enhancing working conditions, improving training quality, and revisiting HRH salaries to mitigate medical student migration [55].

The satisfaction of health professionals with their jobs significantly influences HRH performance and the quality of healthcare services [56,57]. Within this scoping review, dissatisfaction among health professionals across various job aspects is evident [30–32]. Globally, particularly in developing nations, several factors contribute to this decreased job satisfaction among health professionals. Management and leadership, working conditions, staff relations and patient care, responsibility, and workload emerge as key factors linked to job satisfaction [57–59].

Introducing effective interventions has shown promise in enhancing nurses’ job satisfaction and curbing employee turnover [56,57,60]. Additionally, factors such as job security, recognition, respect, financial independence, and competitive salaries are reported in the literature to heighten HRH motivation [61]. Notably, in a national survey of US physicians, intrinsic motivational factors were associated with physicians’ career enjoyment, life satisfaction, and commitment to clinical practice [62].

According to several studies [60,63–65], job satisfaction has been found to correlate with stress levels and the incidence of burnout symptoms within work environments. Stress represents a universal element in the job of healthcare professionals [66]. Chronic occupational stress, heavy workloads, and imbalances between job demands and resources are primary contributors to HRH burnout [67,68].

A meta-analysis of 47 studies revealed that physician burnout increased the risk of medical errors and led to diminished overall care quality and patient satisfaction [69]. However, findings from a review and meta-analysis underscored the effectiveness of cognitive, behavioral, and mindfulness-based approaches in reducing stress levels and burnout among HRH [70]. In our context, studies advocate for enhancing working conditions and providing better financial motivation for public health professionals to mitigate stress and burnout [36,37].

### Limitations

This scoping review bears certain limitations. Firstly, the internal and external validity of the findings might hinder their generalizability to the entire HRH spectrum. The risk of selection bias is a possibility, as only two researchers conducted and agreed upon the database selection. Moreover, the majority of studies included in this review primarily originated from major cities or prominent regions, potentially limiting the diversity and representation of HRH experiences across various settings. Secondly, the predominance of cross-sectional studies focused predominantly on nurses within HRH, along with disparities in sample sizes and variations in the measurement tools used to assess challenges, could impact the broader applicability of the findings.

### Conclusion

This scoping review gathered pertinent documents to offer insights into the prevailing challenges within HRH in Morocco. The identified publications delineated a diverse array of challenges that necessitate robust policy interventions. Despite recent strides in the Moroccan healthcare system, tackling the disparities in HRH distribution between urban and rural areas, augmenting funding, and enhancing the quality of life for HRH remain pivotal. This requires concerted efforts towards strategic planning for staff training, coupled with the implementation of improved HR management practices and enhanced motivation within regional public health institutions.

## Data Availability

All relevant data are within the manuscript and its Supporting Information files.

## References

1. Boniol M, Kunjumen T, Nair TS, Siyam A, Campbell J, Diallo K. The global health workforce stock and distribution in 2020 and 2030: a threat to equity and ‘universal’ health coverage? BMJ Glob Health. 2022;7(6):e009316.

2. Okunogbe A, Bowser D, Gedik G, Naseri S, Abu-Agla A, Safi N. Global Fund financing and human resources for health investments in the Eastern Mediterranean Region. Hum Resour Health. 2020;18(1):48.

3. Campbell J, Dussault G, Buchan J, Pozo-Martin F, Guerra Arias M, Leone C, et al. A Universal Truth: No Health Without A Workforce [Internet]. Recife, Brazil: Geneva, Global Health Workforce Alliance and World Health Organization; 2013 [cited 2023 Sep 23] p. 104. Available from: https://www.who.int/publications/m/item/hrh_universal_truth

4. Crisp N, Chen L. Global Supply of Health Professionals. N Engl J Med. 2014;370(10):950–7.

5. World Health Organization. Global strategy on human resources for health: workforce 2030 [Internet]. Geneva: World Health Organization; 2016 [cited 2023 Nov 7]. 64 p. Available from: https://iris.who.int/handle/10665/250368

6. Belrhiti Z, Van Damme W, Belalia A, Marchal B. The effect of leadership on public service motivation: a multiple embedded case study in Morocco. BMJ Open. 2020;10(1):e033010.

7. Effa E, Arikpo D, Oringanje C, Udo E, Esu E, Sam O, et al. Human resources for health governance and leadership strategies for improving health outcomes in low- and middle-income countries: a narrative review. J Public Health Oxf Engl. 2021;43(Suppl 1):i67–85.

8. Negero MG, Sibbritt D, Dawson A. How can human resources for health interventions contribute to sexual, reproductive, maternal, and newborn healthcare quality across the continuum in low- and lower-middle-income countries? A systematic review. Hum Resour Health. 2021;19(1):54.

9. Rowe AK, Rowe SY, Peters DH, Holloway KA, Chalker J, Ross-Degnan D. Effectiveness of strategies to improve health-care provider practices in low-income and middle-income countries: a systematic review. Lancet Glob Health. 2018;6(11):e1163–75.

10. Sonderegger S, Bennett S, Sriram V, Lalani U, Hariyani S, Roberton T. Visualizing the drivers of an effective health workforce: a detailed, interactive logic model. Hum Resour Health. 2021;19(1):32.

11. Witter S, Hamza MM, Alazemi N, Alluhidan M, Alghaith T, Herbst CH. Human resources for health interventions in high- and middle-income countries: findings of an evidence review. Hum Resour Health. 2020;18(1):43.

12. Aromataris E, Pearson A. The systematic review: an overview. Am J Nurs. 2014;114(3):53–8.

13. Smela B, Toumi M, Świerk K, Gawlik K, Clay E, Boyer L. Systematic literature reviews over the years. J Mark Access Health Policy. 2023;11(1):2244305.

14. Mahdaoui M, Kissani N. Morocco’s Healthcare System: Achievements, Challenges, and Perspectives. Cureus. 2023;15(6):e41143.

15. Chen L, Evans T, Anand S, Boufford JI, Brown H, Chowdhury M, et al. Human resources for health: overcoming the crisis. Lancet Lond Engl. 2004;364(9449):1984–90.

16. Agyeman-Manu K, Ghebreyesus TA, Maait M, Rafila A, Tom L, Lima NT, et al. Prioritising the health and care workforce shortage: protect, invest, together. Lancet Glob Health. 2023;11(8):e1162–4.

17. Arksey H, O’Malley L. Scoping studies: towards a methodological framework. Int J Soc Res Methodol. 2005;8(1):19–32.

18. Levac D, Colquhoun H, O’Brien KK. Scoping studies: advancing the methodology. Implement Sci. 2010;5(1):69.

19. Tricco AC, Lillie E, Zarin W, O’Brien KK, Colquhoun H, Levac D, et al. PRISMA Extension for Scoping Reviews (PRISMAScR): Checklist and Explanation. Ann Intern Med. 2018;169:467–473.

20. Hölbl M, Kompara M, Kamišalić A, Nemec Zlatolas L. A Systematic Review of the Use of Blockchain in Healthcare. Symmetry. 2018;10(10):470.

21. Dankoly US, Vissers D, El Mostafa SB, Ziyyat A, Van Rompaey B, Van Royen P, et al. Perceived barriers, benefits, facilitators, and attitudes of health professionals towards type 2 diabetes management in Oujda, Morocco: a qualitative focus group study. Int J Equity Health. 2023;22:29.

22. El Kirat H, Filali H. La représentation des professionnels de santé mentale sur leurs pratiques à Rabat, Maroc. Santé Publique. 2014;26(3):385–91. French

23. Ministry of Health. Continuing training strategy for health professionals. 2017 p. 68. French

24. Hababa H, Chakiri L, Ouakhzan B, Nairi N. Les enjeux de la formation et du développement du capital humain à l’épreuve de la régionalisation. Int Soc Sci Manag J. 2021 Oct 13;(4). French. Available from: https://revues.imist.ma/index.php/ISSM/article/view/28605

25. Barich F, Chamkal N, Rezzouk B. La formation en soins infirmiers et techniques de santé dans le système licence-master-doctorat au Maroc : analyse des descriptifs de formation, étude analytique descriptive. Rev Francoph Int Rech Infirm. 2019 Dec 1;5(4):100183. French

26. Gouifrane R, Belaaouad S, Benmokhtar S, Radid M. La motivation des infirmiers à la participation aux actions de formation continue : une étude descriptive au niveau du pole des urgences chirurgicales de l’hôpital Ibn-Rochd de Casablanca. Rev Francoph Int Rech Infirm. 2018 Dec 1;4(4):e237–43. French

27. Elkachradi R, Boudallaa I, Hillali M, Kadouri A. Lifelong learning among healthcare professionals in public hospitals: Historical analysis and multiple case studies in Morocco. Bourekkadi S, Kerkeb ML, El Imrani O, Rafalia N, Zubareva O, Khoulji S, et al., editors. E3S Web Conf. 2023;412:01078.

28. Ministry of Health. Universal health coverage in Morocco: challenges for human resources. 2017 p. 33. French

29. Zouag N, Driouchi A, Achehboune A. Labor Health Shortage and Future Prospects for the Medical Workforce in Morocco. MPRA Pap [Internet]. 2015 Apr 10 [cited 2023 Nov 2]; Available from: https://ideas.repec.org//p/pra/mprapa/63547.html

30. El Mouaddib H, Sebbani M, Mansouri A, Adarmouch L, Amine M. Job satisfaction of primary healthcare professionals (public sector): A cross-sectional study in Morocco. Heliyon. 2023;9(9):e20357.

31. Koubri H, Hami H, Soulaymani A, El Kouartey N. Person-centred health services as a new model of the organization of primary healthcare services in Morocco. Journal Xi’an Univ Archit Technol. 2022;14(8):126–34. 10.37896/JXAT14.08/315613

32. Ministry of Health. Study of satisfaction/motivation among health professionals. 2018 p. 73. French

33. El Azizi I, El Goundali. Les barrières liées à l’intégration des résultats probants dans la pratique infirmière : le point de vue d’infirmiers marocains, étude descriptive quantitative. Rev Francoph Int Rech Infirm. 2019;5(4):100181. French

34. Giurgiu DI, Jeoffrion C, Grasset B, Dessomme BK, Moret L, Roquelaure Y, et al. Psychosocial and occupational risk perception among health care workers: a Moroccan multicenter study. BMC Res Notes. 2015;8(1):408.

35. Laraqui O, Manar N, Laraqui S, Boukili M, Ghailan T, Deschamps F, et al. [Job perception and well-being among healthcare workers in Morocco]. Sante Publique. 2017;29(6):887–95. French

36. Fattahi R, Attiya N, FIlali-Zegzouti A, El Haidani A, Bouya S, El Jaafari S, et al. Le burnout parmi le personnel des structures de santé publique de la région de Drâa-Tafilalet au Maroc. Arch Mal Prof Environ. 2023;84(4):101809. French

37. Mansoury O, Chamsi K, Essoli S, Mansouri A, Sebbani M, Adarmouch L, et al. L’épuisement professionnel chez le personnel de santé au centre hospitalier universitaire Mohammed VI de Marrakech – Maroc. Arch Mal Prof Environ. 2023;84(6):101877. French

38. Chtibi H, Ahami A, Azzaoui FZ, Khadmaoui A, Mammad K, Mottier C, et al. Study of Resistance to Stress and Burnout among Public Health Professionals: The Case of Nurses and Physicians at Ibn Sina Hospital in Rabat Morocco. Open J Med Psychol. 2018;7(3):34–46.

39. Endalamaw A, Khatri RB, Erku D, Nigatu F, Zewdie A, Wolka E, et al. Successes and challenges towards improving quality of primary health care services: a scoping review. BMC Health Serv Res. 2023;23(1):893.

40. Jesus TS, Landry MD, Dussault G, Fronteira I. Human resources for health (and rehabilitation): Six Rehab-Workforce Challenges for the century. Hum Resour Health. 2017;15(1):8.

41. Kanchanachitra C, Lindelow M, Johnston T, Hanvoravongchai P, Lorenzo FM, Huong NL, et al. Human resources for health in southeast Asia: shortages, distributional challenges, and international trade in health services. The Lancet. 2011;377(9767):769–81.

42. Su Y, Liu MS, De Silva PV, Østbye T, Jin KZ. Health-related quality of life in Chinese workers: a systematic review and meta-analysis. Glob Health Res Policy. 2021;6:1–17.

43. Benton DC, Shaffer FA. Human Resources for Health 2030 and the regulatory agenda. J Nurs Manag. 2016;24(6):705–7.

44. World Health Assembly 69. Global strategy on human resources for health: workforce 2030. World Health Organization; 2016.

45. Cardarelli R, Koranchelian T. Morocco’s Quest for Stronger and Inclusive Growth. In: Morocco’s Quest for Stronger and Inclusive Growth [Internet]. International Monetary Fund; 2023 [cited 2023 Nov 7]. Available from: https://www.elibrary.imf.org/display/book/9798400225406/9798400225406.xml

46. Fourtassi M, Abda N, Bentata Y, Hajjioui A. Medical education in Morocco: Current situation and future challenges. Med Teach. 2020;42(9):973–9.

47. Massagli TL, Zumsteg JM, Osorio MB. Quality Improvement Education in Residency Training: A Review. Am J Phys Med Rehabil. 2018;97(9):673–8.

48. Tumlinson K, Jaff D, Stilwell B, Onyango DO, Leonard KL. Reforming medical education admission and training in low- and middle-income countries: who gets admitted and why it matters. Hum Resour Health. 2019;17(1):91, s12960-019-0426–9.

49. Micah AE, Solorio J, Stutzman H, Zhao Y, Tsakalos G, Dieleman JL. Development assistance for human resources for health, 1990–2020. Hum Resour Health. 2022;20(1):51.

50. Hachoumi N, Eddabbah M, El Adib AR. Health sciences lifelong learning and professional development in the era of artificial intelligence. Int J Med Inf. 2023;178:105171.

51. O’Malley G, Perdue T, Petracca F. A framework for outcome-level evaluation of in-service training of health care workers. Hum Resour Health. 2013;11(1):50.

52. Frenk J, Chen L, Bhutta ZA, Cohen J, Crisp N, Evans T, et al. Health professionals for a new century: transforming education to strengthen health systems in an interdependent world. The Lancet. 2010;376(9756):1923–58.

53. Tsiachristas A, Wallenburg I, Bond CM, Elliot RF, Busse R, van Exel J, et al. Costs and effects of new professional roles: Evidence from a literature review. Health Policy Amst Neth. 2015;119(9):1176–87.

54. World Health Organization. WHO health workforce support and safeguards list 2023 [Internet]. World Health Organization; 2023 Mar [cited 2023 Nov 8]. Available from: https://www.who.int/publications-detail-redirect/9789240069787

55. Sylla A, El Ouadih S, Barknan K, Hassoune S, Nani S. Migration intention of final year medical students. Eur J Public Health. 2021;31(Supplement_3):ckab165.448.

56. Niskala J, Kanste O, Tomietto M, Miettunen J, Tuomikoski AM, Kyngäs H, et al. Interventions to improve nurses’ job satisfaction: A systematic review and meta-analysis. J Adv Nurs. 2020;76(7):1498–508.

57. Rowan BL, Anjara S, De Brún A, MacDonald S, Kearns EC, Marnane M, et al. The impact of huddles on a multidisciplinary healthcare teams’ work engagement, teamwork and job satisfaction: A systematic review. J Eval Clin Pract. 2022;28(3):382–93.

58. Aloisio LD, Coughlin M, Squires JE. Individual and organizational factors of nurses’ job satisfaction in long-term care: A systematic review. Int J Nurs Stud. 2021;123:104073.

59. Tenaw Z, Siyoum M, Tsegaye B, Werba TB, Bitew ZW. Health Professionals Job Satisfaction and Associated Factors in Ethiopia: A Systematic Review and Meta-analysis. Health Serv Res Manag Epidemiol. 2021;8:23333928211046484.

60. Nowrouzi B, Lightfoot N, Larivière M, Carter L, Rukholm E, Schinke R, et al. Occupational Stress Management and Burnout Interventions in Nursing and Their Implications for Healthy Work Environments: A Literature Review. Workplace Health Saf. 2015;63(7):308–15.

61. Gupta J, Patwa MC, Khuu A, Creanga AA. Approaches to motivate physicians and nurses in low- and middle-income countries: a systematic literature review. Hum Resour Health. 2021;19(1):4.

62. Tak HJ, Curlin FA, Yoon JD. Association of Intrinsic Motivating Factors and Markers of Physician Well-Being: A National Physician Survey. J Gen Intern Med. 2017;32(7):739– 46.

63. Friganović A, Selič P, Ilić B, Sedić B. Stress and burnout syndrome and their associations with coping and job satisfaction in critical care nurses: a literature review. Psychiatr Danub. 2019;31(Suppl 1):21–31.

64. Zhang YY, Zhang C, Han XR, Li W, Wang YL. Determinants of compassion satisfaction, compassion fatigue and burn out in nursing: A correlative meta-analysis. Medicine (Baltimore). 2018;97(26):e11086.

65. Salvagioni DAJ, Melanda FN, Mesas AE, González AD, Gabani FL, Andrade SMD. Physical, psychological and occupational consequences of job burnout: A systematic review of prospective studies. PloS one. 2017;12(10):e0185781.

66. Rink LC, Oyesanya TO, Adair KC, Humphreys JC, Silva SG, Sexton JB. Stressors Among Healthcare Workers: A Summative Content Analysis. Glob Qual Nurs Res. 2023;10:23333936231161127.

67. De Simone S, Vargas M, Servillo G. Organizational strategies to reduce physician burnout: a systematic review and meta-analysis. Aging Clin Exp Res. 2021;33(4):883–94.

68. Molina-Praena J, Ramirez-Baena L, Gómez-Urquiza JL, Cañadas GR, De la Fuente EI, Cañadas-De la Fuente GA. Levels of Burnout and Risk Factors in Medical Area Nurses: A Meta-Analytic Study. Int J Environ Res Public Health. 2018;15(12):2800.

69. Panagioti M, Geraghty K, Johnson J, Zhou A, Panagopoulou E, Chew-Graham C, et al. Association Between Physician Burnout and Patient Safety, Professionalism, and Patient Satisfaction: A Systematic Review and Meta-analysis. JAMA Intern Med. 2018;178(10):1317–31.

70. Regehr C, Glancy D, Pitts A, LeBlanc VR. Interventions to reduce the consequences of stress in physicians: a review and meta-analysis. J Nerv Ment Dis. 2014;202(5):353–9.

